# Parent Race and Communication During Elective Pediatric Surgery Consultations

**DOI:** 10.1101/2025.03.12.25323874

**Authors:** Chenery Lowe, Mary Catherine Beach, Somnath Saha, Anne R. Links, Emily F. Boss

## Abstract

**Importance:** Children of marginalized racial groups have poorer surgical outcomes compared to White children. Clinical communication may contribute to these disparities.

**Objective:** We explore racial differences in parent-clinician communication during initial consultations for elective surgical procedures in children.

**Design:** Cross-sectional study of communication during consultations between surgical clinicians and parents of children (age 2-17) referred for initial evaluation for tonsillectomy/adenoidectomy, hernia repair, and circumcision between 2016 and 2023.

**Setting:** Academically affiliated outpatient clinics in the Baltimore, Maryland region.

**Participants:** Parent-clinician dyads including 22 surgical clinicians and 153 parents.

**Main Outcome(s) and Measure(s):** Visits were audio-recorded and coded using the Roter Interaction Analysis System. Outcomes included visit-level measures (parent versus clinician verbal dominance and patient-centeredness ratio), clinician communication (facilitation/activation statements to encourage parent participation, emotional statements, social chit-chat, and positive affect), and parent communication (emotional statements, social chit-chat, positive affect). We used generalized estimating equations to test associations of parent race with visit communication, accounting for nesting of visits within clinicians and adjusting for parent sociodemographic variables.

**Results:** Of the 153 parents, 63 (41.2%) were Black and 90 (58.8%) were White. Of the 22 clinicians, 14 (63.6%) were White and 8 (36.4%) reported other or multiple ethnicities. In unadjusted models, visits with Black parents had higher clinician verbal dominance but no difference in overall patient-centeredness scores relative to visits with White parents. However, visits with Black parents had fewer clinician chit-chat statements, fewer total parent statements, fewer parent emotional statements, fewer parent chit-chat statements, and lower parent positive affect. After adjusting for parent sociodemographic variables, clinician verbal dominance remained significantly higher, with clinicians making 0.4 more statements per parent statement (95% CI: 0.1 to 0.7).

**Conclusions and Relevance:** In this cohort study of communication during pediatric surgical consultations, parent race was associated with differences in clinician, parent, and interactive visit communication, although some differences appeared to be mediated by sociodemographic factors and most were due to differences in parent rather than clinician communication. Application of patient-centered communication and engagement strategies may help to bridge social distance in pediatric surgical care.

**Key Points:** *Question:* How does parent-clinician communication differ by parent race during initial pediatric surgery consultations?

*Findings:* In this cohort study, consultations with Black parents had higher clinician verbal dominance, less parent emotional expression, and less social chit-chat (both parent and clinician) relative to visits with White parents.

*Meaning:* Our findings reveal opportunities to mitigate racial differences in communication and bridge social distance in pediatric surgical care, including through improved clinician elicitation of patient and family concerns and intentional attempts to build rapport.

## Introduction

In a 2013 meta-analysis, Black children in the U.S. had higher rates of post-operative mortality and morbidity and longer hospital stays relative to White children, even after adjusting for co-morbidities^1^. Multiple factors contribute to this disparity, likely including patient-parent communication. In other settings including adult primary care, clinician bias is associated with less patient-centered communication with patients from marginalized racial/ethnic backgrounds.^4^ In pediatric surgery, this raises concerns about whether parents from marginalized groups are sufficiently able to discuss their concerns about the risks, benefits, and options when making surgical decisions. There is limited evidence about racial differences in communication during pediatric surgical consultations.

Our objective was to explore racial differences in communication during initial consultations for three common referral indications: tonsillectomy/adenoidectomy, hernia repair, and circumcision. We hypothesized that visits with Black parents would have higher clinician verbal dominance and lower patient-centeredness relative to visits with White parents. We explored race’s associations with specific aspects of clinician communication (facilitation/activation statements, social chit-chat, emotional statements, and global affect) and parent communication (social chit-chat, emotional statements, and global affect).

## Methods

We followed the STROBE guidelines for reporting observational cohort studies ^5^.

### Participants and Data Collection

Data were collected at three outpatient otolaryngology clinics, one urology clinic, and one general surgery outpatient clinic at Johns Hopkins Medicine (2016-2023), as part of a study evaluating communication, shared decision-making, and bias in pediatric elective surgery consultations. Clinicians were eligible if they were an attending physician or advanced practice provider who regularly evaluated children for tonsillectomy or sleep-disordered breathing, hernia repair, or circumcision. Statements from supporting clinicians (e.g., residents, other clinical staff) were included in recordings and coding, but demographics for these clinicians was not available. Parents were eligible if their child underwent initial evaluation for sleep-disordered breathing or tonsillectomy, hernia, or circumcision. Parents were identified via electronic health records and contacted before their visit. Participants completed written informed consent before engaging in study activities, then completed demographic questionnaires. Consultations were audio-recorded. Johns Hopkins School of Medicine Institutional Review Board approved all procedures and was the IRB of record for the study (IRB00259996).

### Parent, child, and clinician sociodemographic variables

The parent sociodemographic questionnaire included self-reported questions about parent age, parent gender, parent race, parent ethnicity, annual household income, parent education, insurance type, child procedure, child gender, child age, child race, and child ethnicity. Clinicians reported their gender, race, ethnicity, and years of experience (Table 2). Race response options included: 1) American Indian or Alaska Native, 2) Asian, 3) Black or African American, 4) Native Hawaiian or Other Pacific Islander, 5) White, 6) Other. Ethnicity options included: 1) No, not of Hispanic, Latino, or Spanish origin, 2) Yes, Mexican, Mexican American, Chicano, 3) Yes, Cuban, and 4) Yes, another Hispanic, Latino, or Spanish origin. Respondents could select multiple options for race and ethnicity. Those selecting “Other” in response to race or “Yes, another Hispanic, Latino, or Spanish origin” in response to ethnicity could specify in a free response box. For this analysis, parents were considered to be non-Hispanic/Latino White (referred to as White) if their selected race was White and ethnicity as “No, not of Hispanic, Latino, or Spanish origin” and Black if they selected their race as Black or African American.

**Table 1.**
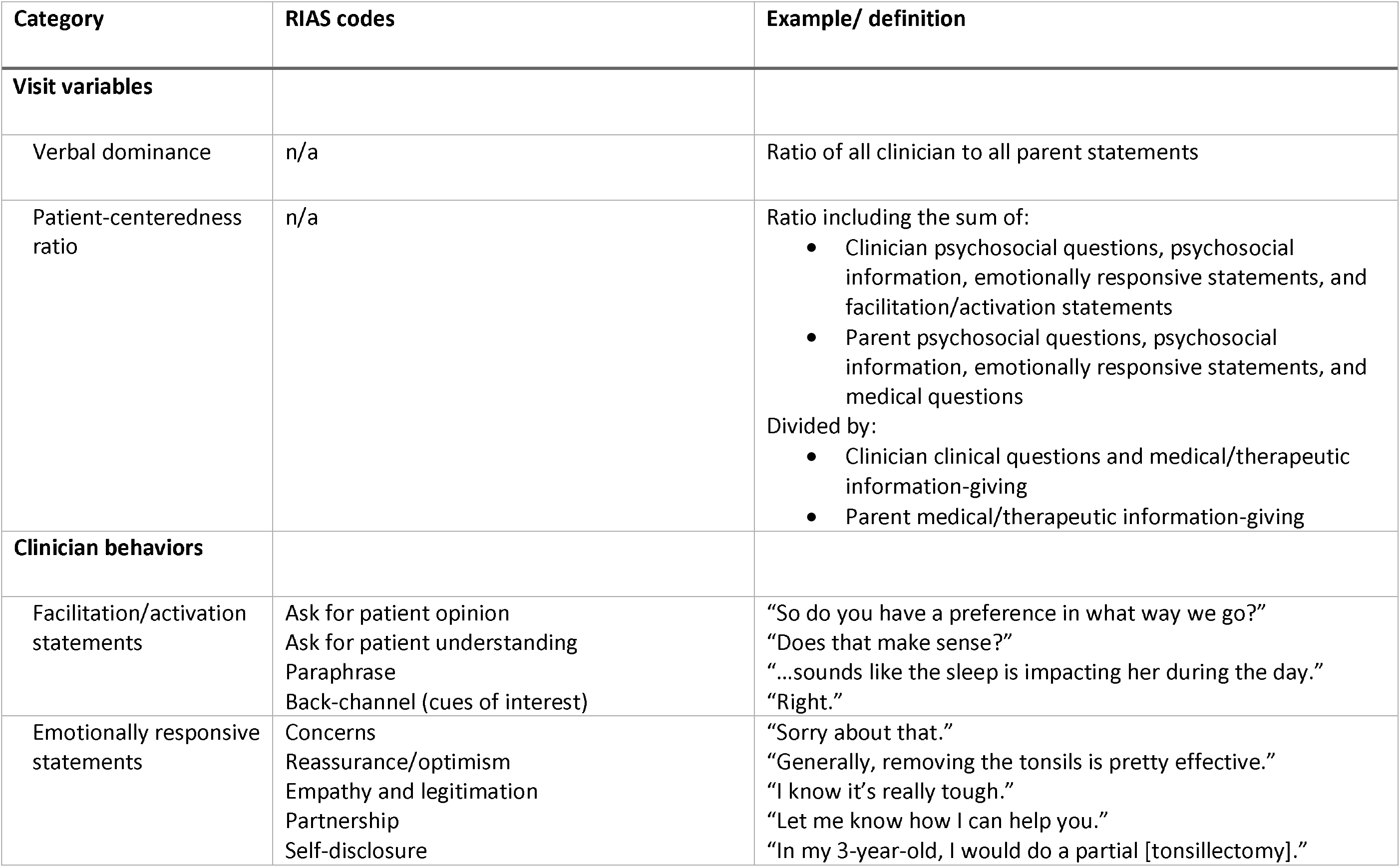
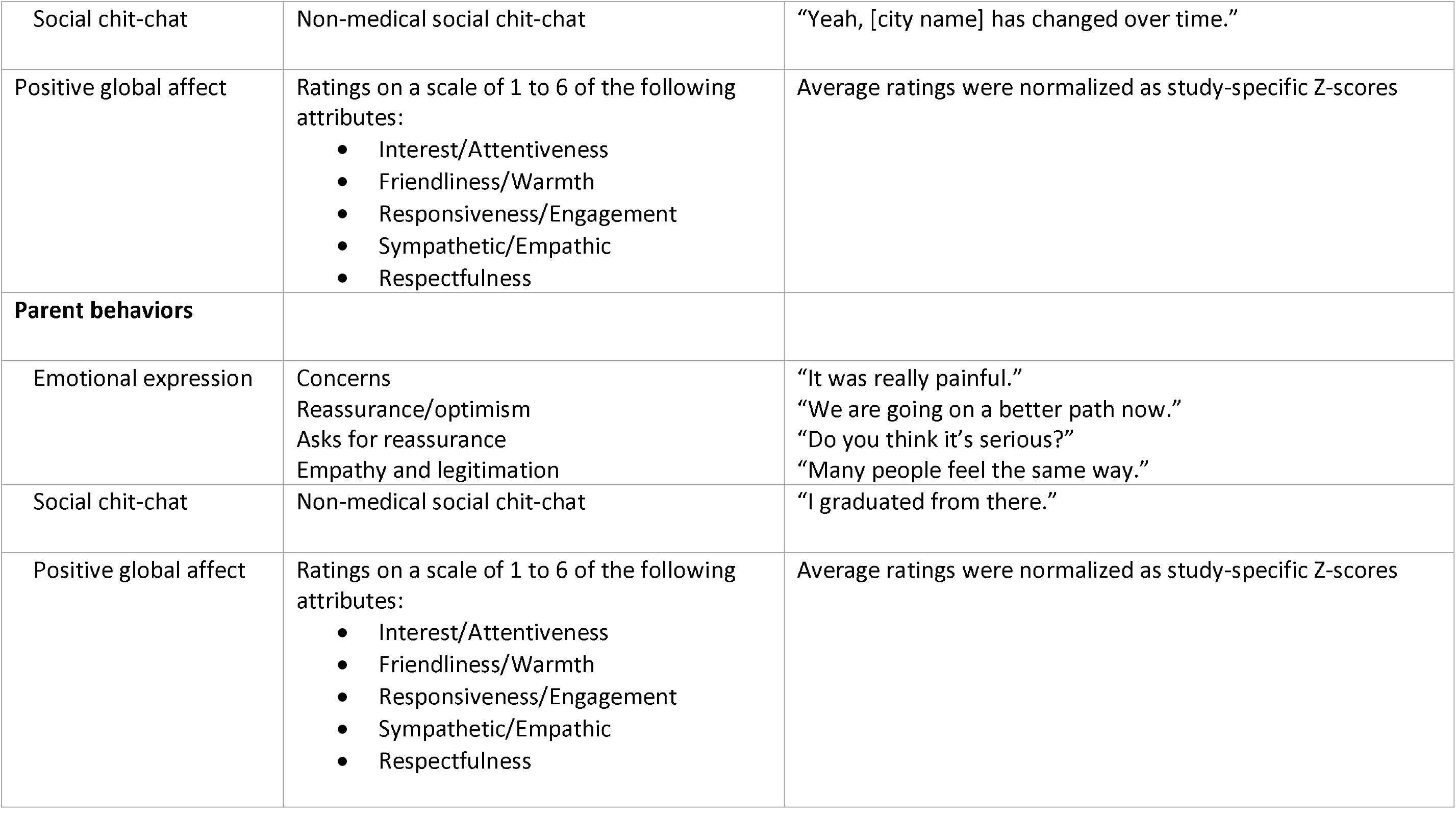
Parent and clinician communication behaviors and examples.

| Category | RIAS codes | Example/ definition |
| --- | --- | --- |
| **Visit variables** |  |  |
| Verbal dominance | n/a | Ratio of all clinician to all parent statements |
| Patient-centeredness ratio | n/a | Ratio including the sum of:<br>• Clinician psychosocial questions, psychosocial information, emotionally responsive statements, and facilitation/activation statements<br>• Parent psychosocial questions, psychosocial information, emotionally responsive statements, and medical questions<br>Divided by:<br>• Clinician clinical questions and medical/therapeutic information-giving<br>• Parent medical/therapeutic information-giving |
| **Clinician behaviors** |  |  |
| Facilitation/activation statements | Ask for patient opinion<br>Ask for patient understanding<br>Paraphrase<br>Back-channel (cues of interest) | "So do you have a preference in what way we go?"<br>"Does that make sense?"<br>"...sounds like the sleep is impacting her during the day."<br>"Right." |
| Emotionally responsive statements | Concerns<br>Reassurance/optimism<br>Empathy and legitimation<br>Partnership<br>Self-disclosure | "Sorry about that."<br>"Generally, removing the tonsils is pretty effective."<br>"I know it's really tough."<br>"Let me know how I can help you."<br>"In my 3-year-old, I would do a partial [tonsillectomy]." |
| Social chit-chat | Non-medical social chit-chat | "Yeah, [city name] has changed over time." |
| Positive global affect | Ratings on a scale of 1 to 6 of the following attributes:<br>• Interest/Attentiveness<br>• Friendliness/Warmth<br>• Responsiveness/Engagement<br>• Sympathetic/Empathic<br>• Respectfulness | Average ratings were normalized as study-specific Z-scores |
| **Parent behaviors** |  |  |
| Emotional expression | Concerns<br>Reassurance/optimism<br>Asks for reassurance<br>Empathy and legitimation | "It was really painful."<br>"We are going on a better path now."<br>"Do you think it's serious?"<br>"Many people feel the same way." |
| Social chit-chat | Non-medical social chit-chat | "I graduated from there." |
| Positive global affect | Ratings on a scale of 1 to 6 of the following attributes:<br>• Interest/Attentiveness<br>• Friendliness/Warmth<br>• Responsiveness/Engagement<br>• Sympathetic/Empathic<br>• Respectfulness | Average ratings were normalized as study-specific Z-scores |

**Table 2.**
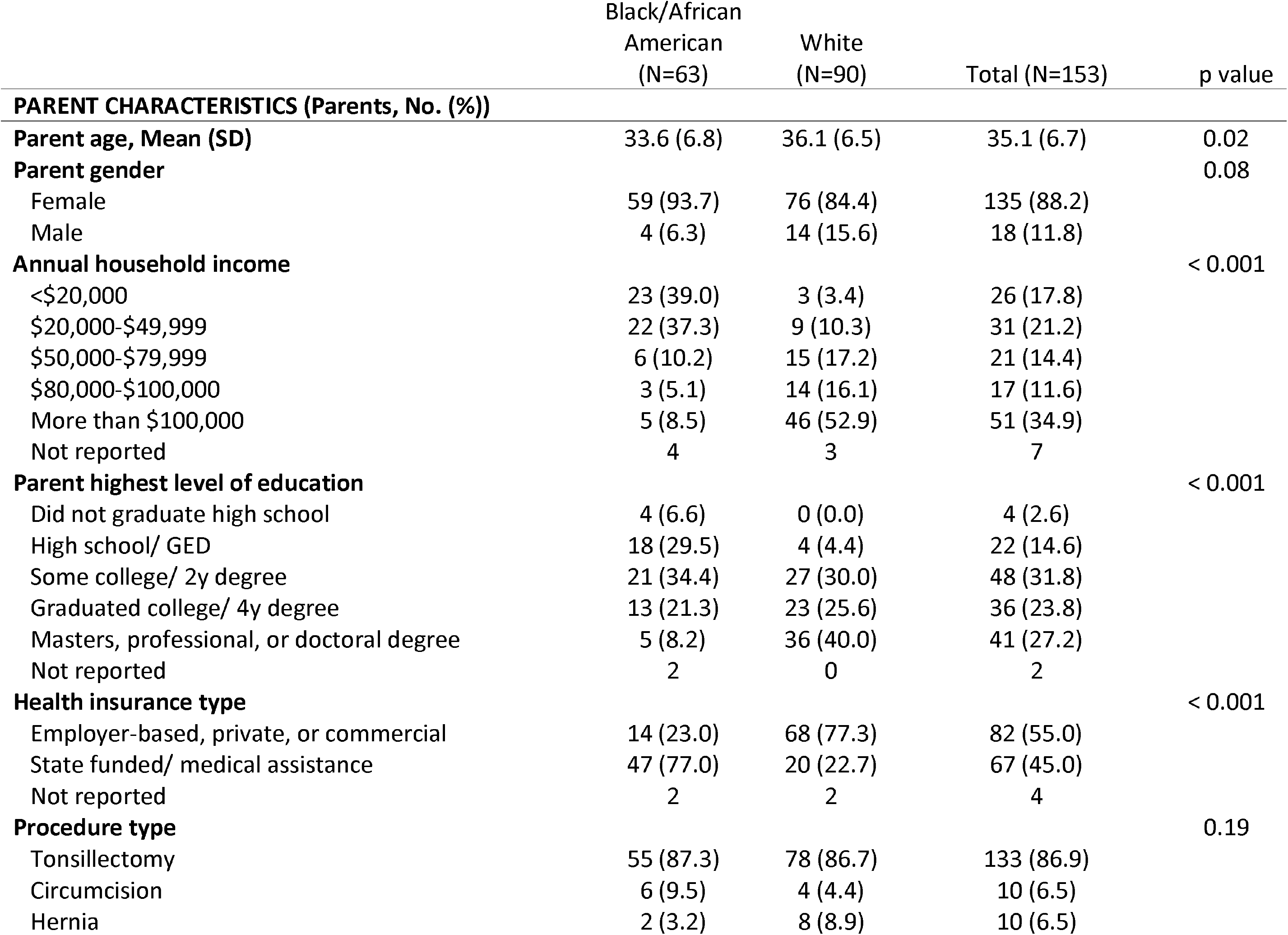
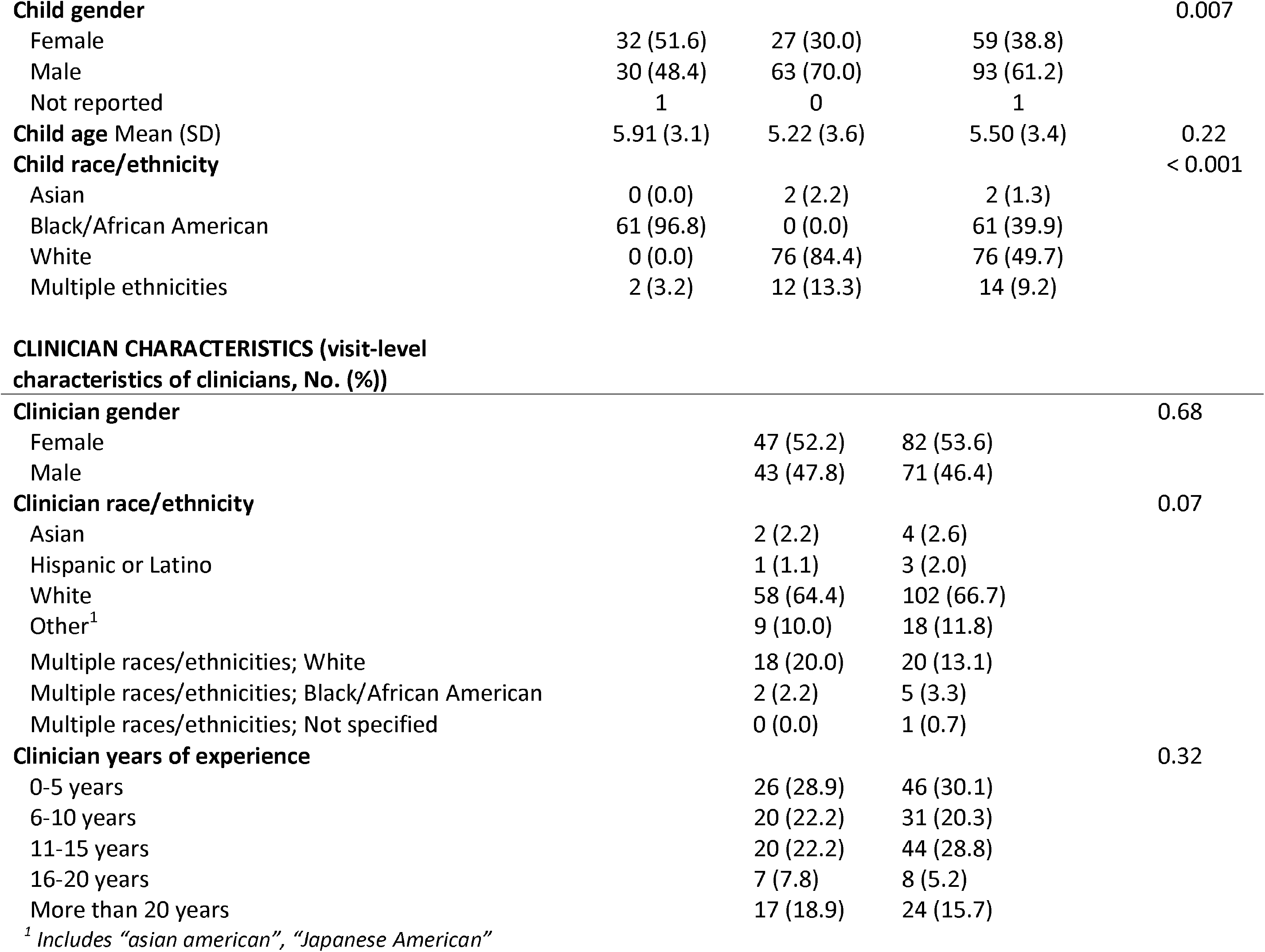
Visit characteristics.

### Communication Coding

A single coder with more than 10 years of experience coded visit recordings using the Roter Interaction Analysis System (RIAS), a widely-used quantitative communication coding system developed for medical interactions^6^. The coder did not view participants’ surveys but was not blinded to demographic information that could be determined from listening to the recording. Coder drift was assessed by recoding and calculating intra-coder reliability for a random sample of visits, but reliability data was not available at the time of this interim analysis. RIAS coding categorizes each unit of clinician or patient expression that conveys a complete thought into one of 37 mutually exclusive and exhaustive codes reflecting biomedically- and socioemotionally-focused statements.^6^ Codes were combined to produce categories described below, with definitions and examples in Table 1.

#### Visit-level measures

Coded statements were combined to calculate verbal dominance, the ratio of all clinician statements to all patient statements during the visit. Codes were also combined to calculate the patient-centeredness ratio, which reflects the balance between socioemotional and biomedical exchange, with higher values indicating a stronger socioemotional focus.

#### Clinician communication behaviors

Codes were combined to capture clinicians’ facilitation/activation statements, emotionally responsive statements, and social chit-chat. These categories are subsets of socioemotionally focused communication. When more than one clinician spoke during a visit, coding distinguished between the primary and supporting clinicians, typically an advanced practice provider or trainee. To ensure that analyses captured the patient’s entire experience of the consultation, the RIAS outcome variables reflect communication with both the primary and supporting clinicians.

#### Parent communication behaviors

RIAS codes were combined to create categories reflecting parent emotional expression and social chit-chat. Coding did not distinguish between the speakers when more than one adult caregiver was present during a visit.

#### Affect ratings

After coding each visit, the coder rated clinician and parent affect during the visit using a Likert scale ranging from one to six. Positive clinician and parent affect were each determined by calculating the average of global ratings across five dimensions: interest/attentiveness, friendliness/warmth, responsiveness/engagement, sympathetic/empathetic, and respectfulness. Due to the potential for coder drift, these ratings were normalized as Z-scores based on the mean affect ratings from each study data collection period.

### Statistical Analysis

Analyses included parents who identified as Black or White. Because parents of other races and ethnicities represented a small proportion of the study population (n=29/182, 15.9%), we lacked sufficient sample size to conduct comparative analyses by parent race. Bivariate analyses assessed the unadjusted association of parent race with the outcomes listed in Table 1. Differences in sociodemographic characteristics by parent race were assessed using ANOVA for continuous variables (parent, child, and clinician age) and chi-square tests for categorical variables. We used generalized estimating equations with an exchangeable correlation structure to evaluate the association of parent race with each communication behavior, accounting for clustering within clinicians. Since race can influence outcomes both directly and indirectly (e.g., through association with other forms or privilege or disadvantage), we assessed unadjusted models and models that adjusted for parent age, gender, insurance, and parent education (trichotomized as “High school or less,” “Some college or 2-year degree”, or “4-year degree or any postgraduate education”). Because annual household income and health insurance type were correlated, only insurance type was included as a model covariate because it was theorized to be more closely related to health care access. Both measures were correlated with parent race. Missing data were considered to be missing completely at random. Reasons for missing data were generally unrelated to participant or visit characteristics and include inability to code the visit due to poor quality audio recording or need for additional processing, technical issues with the recording, or multiple children being evaluated during the visit (n=13/166, 7.8%) as well as non-completion of survey measures (Table 2). Analyses were conducted using R Statistical Software (Version 4.3.1) ^7^.

## Results

### Participant characteristics

#### Parent and child characteristics

Of 182 enrolled parent-patient dyads enrolled in the study, 153 (84.0%) were either Black or White. To facilitate statistical comparisons, visits with the 153 parents who identified as Black (n=63, 41%) or White (n=90, 59%) were included in analysis. The average parent age was 33.6 years for Black parents and 36.1 years for White parents. Most parents were female (93.7% Black parents, 84.4% of White parents). Additional demographics are shown in Table 2. Relative to Black parents, White parents were older, had higher annual family income, and more education. White parents were more likely to have employer-based, private, or commercial health insurance rather than state-funded or medical assistance insurance and had a higher proportion of male relative to female children being evaluated for surgery.

#### Clinician characteristics

Clinicians (n=22) were predominantly female (54.5%). Thirteen clinicians identified as White (59.1%), 2 (9.1%) were Asian, 1 (4.5%) Hispanic or Latino, 3 (13.6%) as another race or ethnicity, and 3 (13.6%) as multiple races/ethnicities that included Black/African American for one and White for another. Because data were collected over multiple years, clinician age and years of experience varied. Table 2 shows the characteristics of clinicians involved in the 153 consultations. Clinicians conducted study visits with an average of 7.0 enrolled families each (median: 3, sd: 7.9, range: 1 to 24 visits per clinician). A supporting clinician such as a medical resident or nurse participated in 99 (64.7%) of visits. Typically, the supporting clinician would see the family unsupervised before the attending joined the visit.

### Visit-level communication

There was no difference in total visit length by parent race (White parents mean: 18.0 minutes, s.d. 6.9 minutes; Black parents mean 17.2 minutes, s.d. 6.7 minutes). Clinicians’ verbal dominance was lower during visits with White parents relative to Black parents, indicating that clinician statements represented a larger share of the dialogue relative to parent statements (Table 3). This association was significant in the unadjusted model (Model 1), with clinicians making 0.4 (95% CI: 0.1 to 0.8) more statements per parent statement in visits with Black parents. This association was significant after adjusting for parent age, gender, and insurance type (Model 2, β=0.5, 95% CI: 0.1 to 0.8), and after additionally adjusting for parent education (Model 3, β=0.4, 95% CI: 0.1 to 0.7). Models excluding dialogue between the supporting clinician and the parent(s) found similar unadjusted and adjusted relationships between parent race and verbal dominance (eTable 1). Since differences in verbal dominance may reflect clinicians talking more, parents talking less, or both, we explored the effects of parent race on total clinician and parent statements separately. The unadjusted model (Table 3, Model 1) indicates that Black parents made 26.6 (95% CI: 46.0 to 7.2) fewer total statements. While models that adjusted for parent demographics (Models 2 and 3) show that visits with Black parents had fewer parent statements and more clinician statements, none of these individual differences was statistically significant at the P<0.05 level. There were no differences by parent race for patient-centeredness (Table 3).

**Table 3.**
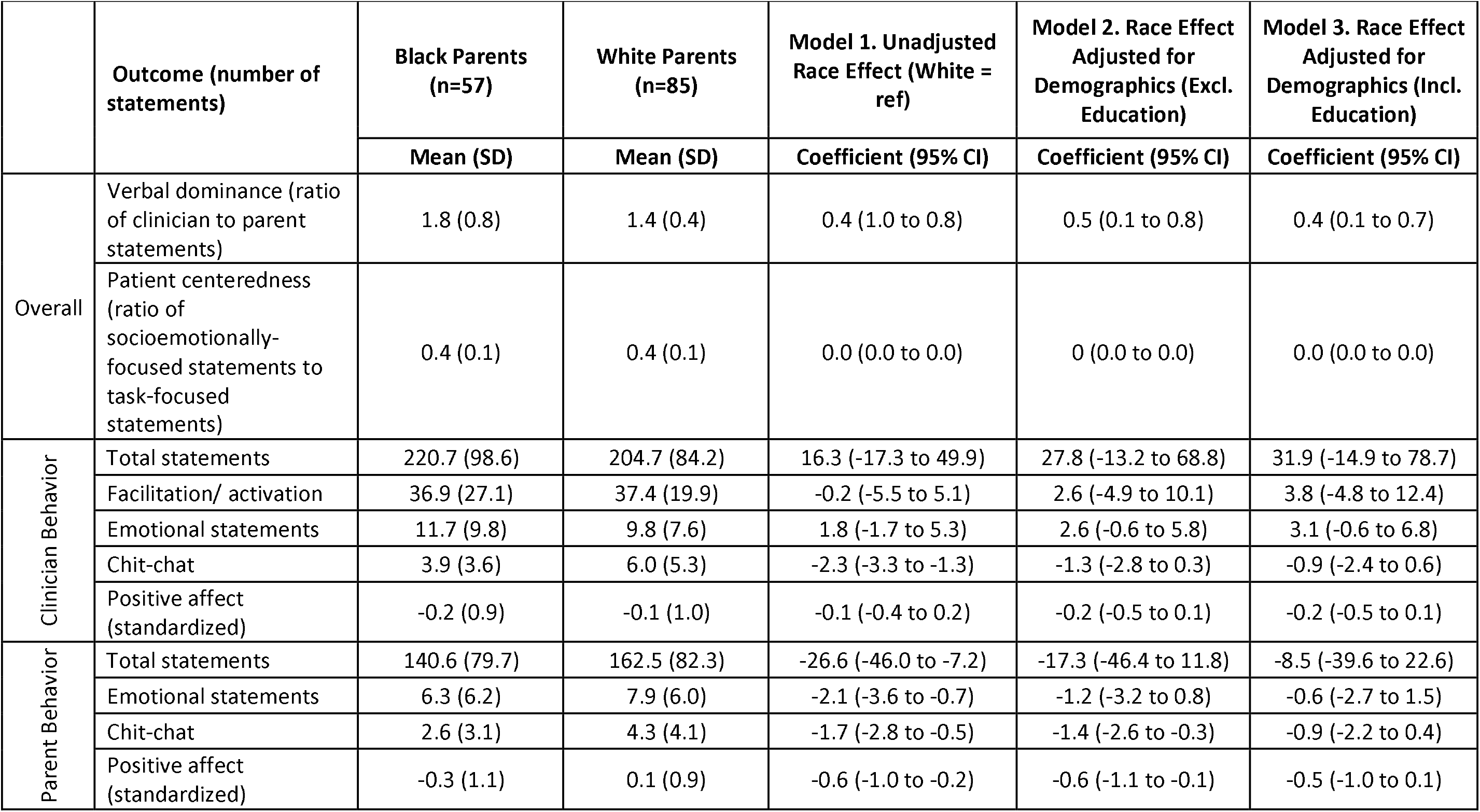
Unadjusted and Adjusted Effects of Parent Race on Communication During Consultations.

|  | Outcome (number of statements) | Black Parents (n=57) | White Parents (n=85) | Model 1. Unadjusted Race Effect (White = ref) | Model 2. Race Effect Adjusted for Demographics (Excl. Education) | Model 3. Race Effect Adjusted for Demographics (Incl. Education) |
| --- | --- | --- | --- | --- | --- | --- |
|  |  | Mean (SD) | Mean (SD) | Coefficient (95% CI) | Coefficient (95% CI) | Coefficient (95% CI) |
| Overall | Verbal dominance (ratio of clinician to parent statements) | 1.8 (0.8) | 1.4 (0.4) | 0.4 (1.0 to 0.8) | 0.5 (0.1 to 0.8) | 0.4 (0.1 to 0.7) |
|  | Patient centeredness (ratio of socioemotionally-focused statements to task-focused statements) | 0.4 (0.1) | 0.4 (0.1) | 0.0 (0.0 to 0.0) | 0 (0.0 to 0.0) | 0.0 (0.0 to 0.0) |
| Clinician Behavior | Total statements | 220.7 (98.6) | 204.7 (84.2) | 16.3 (-17.3 to 49.9) | 27.8 (-13.2 to 68.8) | 31.9 (-14.9 to 78.7) |
|  | Facilitation/ activation | 36.9 (27.1) | 37.4 (19.9) | -0.2 (-5.5 to 5.1) | 2.6 (-4.9 to 10.1) | 3.8 (-4.8 to 12.4) |
|  | Emotional statements | 11.7 (9.8) | 9.8 (7.6) | 1.8 (-1.7 to 5.3) | 2.6 (-0.6 to 5.8) | 3.1 (-0.6 to 6.8) |
|  | Chit-chat | 3.9 (3.6) | 6.0 (5.3) | -2.3 (-3.3 to -1.3) | -1.3 (-2.8 to 0.3) | -0.9 (-2.4 to 0.6) |
|  | Positive affect (standardized) | -0.2 (0.9) | -0.1 (1.0) | -0.1 (-0.4 to 0.2) | -0.2 (-0.5 to 0.1) | -0.2 (-0.5 to 0.1) |
| Parent Behavior | Total statements | 140.6 (79.7) | 162.5 (82.3) | -26.6 (-46.0 to -7.2) | -17.3 (-46.4 to 11.8) | -8.5 (-39.6 to 22.6) |
|  | Emotional statements | 6.3 (6.2) | 7.9 (6.0) | -2.1 (-3.6 to -0.7) | -1.2 (-3.2 to 0.8) | -0.6 (-2.7 to 1.5) |
|  | Chit-chat | 2.6 (3.1) | 4.3 (4.1) | -1.7 (-2.8 to -0.5) | -1.4 (-2.6 to -0.3) | -0.9 (-2.2 to 0.4) |
|  | Positive affect (standardized) | -0.3 (1.1) | 0.1 (0.9) | -0.6 (-1.0 to -0.2) | -0.6 (-1.1 to -0.1) | -0.5 (-1.0 to 0.1) |

### Clinician communication

Clinician social chit-chat was lower during visits with Black relative to White parents, with clinicians making 2.3 (95% CI: 3.3 to 1.3) fewer chit-chat statements per consultation with Black parents in the unadjusted model (Table 3, Model 1). After adjusting for parent characteristics (Models 2 and 3) there was no statistically significant difference in clinician chit-chat statements by parent race. There were no differences in clinician facilitation/activation statements and emotionally responsive statements (Table 3).

### Parent communication

Black parents made 1.7 (95% CI: 2.8, 0.55) fewer chit-chat statements than White parents in the unadjusted model (Table 3, Model 1) and in a model that adjusted for parent age, gender, and insurance (Model 2, β=−1.4, 95% CI: −2.6 to −0.3). However, racial difference was no longer significant after adjusting for parent education (Model 3, β=−0.9, 95% CI: −2.2, 0.4). Black parents made 2.1 (95%CI: 3.6 to 0.7) fewer emotional statements than White parents in the unadjusted model (Table 3, Model 1). However, after adjusting for parent demographics, the relationship between parent race and emotional statements was not statistically significant (Models 2 and 3).

### Clinician and parent affect

Internal consistency across positive affect categories was adequate for both parent (standardized Cronbach’s alpha = 0.8, 95% CI: 0.81 to 0.89) and clinician (standardized Cronbach’s alpha = 0.82, 95% CI: 0.75 to 0.87) ratings. White parents had higher positive affect ratings compared to Black parents in the unadjusted model (Table 3, Model 1) and in the model that adjusted for parent age, gender, and insurance type (Model 2). This effect was attenuated after adjusting for parent education (Model 3). There were no racial differences in clinician positive affect in adjusted or unadjusted models (Table 3).

## Discussion

While parent race was not associated with many of the communication behaviors assessed, visits with Black parents had higher clinician verbal dominance and other differences in parents’ emotional expression and positive affect as well as parents’ and clinicians’ social chit-chat. While most of these racial differences are due to differences in parent rather than clinician statements, they nonetheless suggest racial differences in the interpersonal care experienced by parents. Collectively, these associations are consistent with a pattern of social distance between predominantly White clinicians and Black relative to White parents, which may contribute to disparities in care quality and outcomes.

Verbal dominance was lower in visits with White relative to Black parents. Adjusting for parent demographics did not reduce this difference. Differences in verbal dominance ratios appear to be primarily due to Black parents making fewer total statements in the unadjusted model. Higher verbal dominance during visits with patients of marginalized racial and ethnic backgrounds has been previously observed in other settings including primary care and oncology ^8–13^, although the finding is not consistent across studies and settings ^14^. In primary care, verbal dominance and the similar metric of clinician-patient talk-time ratio have been previously associated with clinician’s scores on the Race Implicit Association Test ^15^ – a measure of implicit racial bias^4,16^ – and patients’ perceived past discrimination,^16^ and decreased adherence to recommendations.^16^ Future research should further explore the role of implicit bias and the effects of parent participation on decisions, care experiences, and clinical outcomes during and following pre-surgical consultations.

Parent emotional expression was more frequent in visits with White relative to Black parents in the unadjusted model. This may reflect differences in parents’ spontaneous expression of emotion, clinicians’ effectiveness at eliciting parent emotions, or combined parent and clinician factors. In a study that analyzed communication between predominantly White clinicians interacting with HIV patients, Black patients were less likely to spontaneously discuss their emotions than White patients, but clinicians were also more likely to block emotional expression and less likely to respond in ways that provided space for further discussion of the emotional issue.^17^ Inadequate emotional exchange is concerning in initial surgical consultations because clinicians may miss opportunities to respond to parent concerns that are relevant to treatment decision-making. Future research to elucidate the impact of unequal emotional expression includes exploring the content of emotional exchange – using methods such as conversation analysis^18^, Verona Coding^19^, or qualitative approaches – and examining how emotional statements are associated with parent-reported and clinical outcomes.

Parent and clinician chit-chat were less frequent in visits with Black relative to White parents. Black parents also had lower observer-reported positive affect than White parents, potentially signaling lower engagement or satisfaction. Both findings suggest differences in rapport-building and the quality of the patient-clinician relationship. While there is limited research on non-medical social talk, we reason that it strengthens relationships by conveying warmth and appreciation for the conversation partner, encouraging engagement and disclosure. Chit-chat can potentially help clinicians to see patients as unique individuals rather than as members of groups, which may mitigate implicit bias and stereotyping ^20,21^.

Education may have both direct and indirect relationships with communication. Education can contribute to higher health literacy and empowerment, resulting in more active participation. More education may also influence clinician communication implicitly by signaling less social distance between parents and clinicians. White parents reported higher educational attainment than Black parents in our study. However, racial differences in verbal dominance remained in regression models that adjusted for parent education, suggesting that this dynamic is not solely influenced by individual differences in education-linked factors such as patient understanding and empowerment. By contrast, adjusting for parent education attenuated the strength of association between parent race and the outcomes of chit-chat and parent positive affect. These findings collectively suggest that structural forces such as racial disparities in educational access interact with interpersonal dynamics to produce communication differences in this setting. Thus, disparities in parent characteristics that are linked to education – such as knowledge, empowerment, and health literacy – should be taken into account when developing mitigation strategies.

### Limitations

The study population included participants from a limited number of outpatient clinics from a single health system in the Baltimore, Maryland region. Moreover, while multiple indications are represented, tonsillectomy/adenoidectomy accounted for more than 80% of analyzed visits. Our findings may therefore not be generalizable to all contexts or indications. This analysis only assessed communication with Black and White parents due to the relatively low representation of parents of other races or ethnicities in this study. Due to the small number of clinicians and their limited racial and ethnic diversity, we were unable to examine associations with clinician race/ethnicity and racial/ethnic concordance between clinicians and patients. We also had limited information about supporting clinicians, making it difficult to assess how their characteristics influenced visit communication (although their communication was included in the analysis.) Additionally, as an analytic method, RIAS captures general categories of medical task-focused and socio-emotional communication but is more limited in measuring contextual and linguistic aspects of clinical communication^6^.

### Implications for practice and future research

Avenues for future research include examining the relationship between visit communication, parent-reported outcomes, and clinical outcomes; exploring how clinician bias influences the relationship between patient race and communication; correlating quantitative findings with qualitative analysis of communication; and further examining how communication with multiple clinicians during consultations influences the quality and equity of care. These results also suggest intervention approaches to promote equitable communication in this setting. Clinician behaviors that may help to encourage parent participation include using eliciting parent perspectives, asking open-ended questions, responding to parent emotions, and engaging in rapport-building social talk. Patient activation interventions that enhance parent empowerment and knowledge may also be valuable.

## Conclusions

In this cohort study of parent communication during pediatric surgical consultations, visits with Black parents had higher verbal dominance, less social chit-chat, fewer parent emotional statements, and lower positive parent affect compared to visits with White parents. Verbal dominance was higher in visits with Black parents, regardless of adjustment for parent sociodemographic factors. These findings signal opportunities for clinicians to encourage parent participation in decision-making for elective pediatric surgical procedures.

## Supporting information

eTable1

## Data sharing statement

Data is not shared.

## Acknowledgements

We thank Michele Massa and Debra Roter for their assistance related to RIAS data coding and management.

Emily Boss and Chenery Lowe had full access to all the data in the study and take responsibility for the integrity of the data and the accuracy of the data analysis.

Research reported in this study was supported by the National Heart, Lung, and Blood Institute (NHLBI) of the National Institutes of Health (NIH) under award R01HL166504. Dr Lowe is supported by a grant from the National Human Genome Research Institute (T32HG008953; the Stanford Training Program in the Ethical, Legal and Social Implications Research Program).

The funders had no role in the design and conduct of the study; collection, management, analysis, and interpretation of the data; preparation, review, or approval of the manuscript; and decision to submit the manuscript for publication.

The content is solely the responsibility of the authors and does not necessarily represent the official views of the NHLBI or the NHGRI of the NIH.

The authors have no potential conflicts of interest to disclose.

