## Supplementary material for "Parent Race and Communication During Elective Pediatric Surgery Consultations": eTable1

### Online-Only Table

**eTable 1. Unadjusted and Adjusted Effects of Parent Race on Communication With Primary Clinicians Only**

|  | Outcome | **Mean (SD) for Black Parents (n=57)^1^** | **Mean (SD) for White Parents (n=85)** | **Model 1. Unadjusted Race Effect (White = ref)** | **Model 2. Race Effect Adjusted for Demographics (Excl. Education)** | **Model 3. Race Effect Adjusted for Demographics (Incl. Education)** |
| --- | --- | --- | --- | --- | --- | --- |
|  |  | **Mean (SD)** | **Mean (SD)** | **Coefficient (95% CI)** | **Coefficient (95% CI)** | **Coefficient (95% CI)** |
| Overall | Verbal dominance | 1.81 (0.75) | 1.39 (0.45) | 0.4 (0.08 to 0.71) | 0.41 (0.1 to 0.73) | 0.36 (0.06 to 0.66) |
|  | Patient centeredness | 0.37 (0.14) | 0.37 (0.12) | 0 (-0.04 to 0.03) | 0.01 (-0.03 to 0.05) | 0.02 (-0.01 to 0.06) |
| Clinician Behavior | Total clinician statements | 184.32 (80.05) | 154.87 (68.86) | 14.46 (-18.42 to 47.34) | 14.17 (-18.28 to 46.63) | 15.43 (-15.66 to 46.53) |
|  | Clinician facilitation/ activation statements | 28.09 (14.27) | 26.89 (14.25) | -0.35 (-4.04 to 3.34) | -0.5 (-5.69 to 4.68) | -0.81 (-5.16 to 3.54) |
|  | Clinician emotional statements | 9.91 (9.39) | 7.09 (5.71) | 1.67 (-1.55 to 4.88) | 1.96 (-0.86 to 4.79) | 2.27 (-0.95 to 5.49) |
|  | Clinician chit-chat statements | 3.04 (2.58) | 3.84 (3.69) | -0.69 (-1.68 to 0.3) | -0.4 (-1.18 to 0.39) | -0.32 (-1.18 to 0.55) |
| Parent Behavior | Total parent statements | 113.35 (56.22) | 118.76 (53.3) | -4.79 (-22.41 to 12.83) | -10.24 (-24.38 to 3.89) | -2.48 (-22.76 to 17.81) |
|  | Parent emotional statements | 5.11 (5.49) | 6.16 (4.5) | -0.78 (-1.88 to 0.32) | -1.04 (-2.65 to 0.58) | -2.53 (-5.99 to 0.93) |
|  | Parent chit-chat statements | 2.32 (3.05) | 2.87 (2.88) | -0.56 (-1.39 to 0.28) | -0.27 (-0.94 to 0.4) | 0.02 (-0.87 to 0.9) |
